# Artificial Intelligence in Healthcare: 2024 Year in Review

**DOI:** 10.1101/2025.02.26.25322978

**Authors:** Raghav Awasthi, Sai Prasad Ramachandran, Shreya Mishra, Dwarikanath Mahapatra, Hajra Arshad, Aarit Atreja, Anirban Bhattacharyya, Atharva Bhattad, Nishant Singh, Jacek B. Cywinski, Ashish K. Khanna, Kamal Maheshwari, Chintan Dave, Avneesh Khare, Francis A. Papay, Piyush Mathur

## Abstract

**Background:** With over a thousand FDA-approved artificial intelligence/machine learning-enabled medical devices, research and publications is maturing from focusing on the development and internal validation of models to the external validation of models and implementation trials. Foundation models, especially Large Language Models, have spurred additional aspects of AI research related to healthcare, especially with the use of text-based data to address healthcare education and administrative tasks related to patient care.

**Methods:** We performed a PubMed search using the terms “machine learning” or “artificial intelligence” and “2024,” restricted to English language and human subject research on January 1, 2025. Utilizing a deep learning-based approach, we assessed the maturity of publications. Following this, we manually annotated the healthcare specialty, data utilized, and models employed for the identified mature articles. Subsequently, empirical data analysis was performed to elucidate trends and statistics. We also performed a detailed analysis of the distribution of foundation model-based publications amongst the healthcare specialties.

**Results:** For the year 2024, the PubMed search yielded 28,180 articles, of which 1,693 were classified as mature using a BERT model. Following exclusions, 1,551 articles were selected for the final data analysis. Amongst these, the highest number of articles in each specialty originated from Imaging (407), Head and Neck (127), and General (122). The analysis of data types revealed that image data (903 [57.0%]) was still the predominant data type, but the use of text data (525 [33.1%]) had substantially increased. Additionally, we also found that LLMs (479) and AI General (448) category models have overtaken deep learning models (372) in healthcare AI research. For LLM-related publications, we are seeing increasing trends in research related to healthcare education and administrative tasks.

**Conclusion:** With the introduction of foundation models, healthcare research trends are changing. The adoption of LLMs and text data types amongst various healthcare specialties, especially for education and administrative tasks, is unlocking new potential for AI applications in healthcare.

## INTRODUCTION

Artificial intelligence has seen a substantial change in research and publication trends since the introduction of ChatGPT [1]. We saw a similar shift in research trends related to AI in healthcare in 2023 [2]. In the past, deep learning models using image-based data were the primary method used in research and publications related to AI in healthcare [3]. These publications were predominantly in the fields of Imaging (Radiology), Oncology, Pathology, and Ophthalmology, amongst others [3]. With the introduction of Large Language Models(LLMs), we saw a shift towards using text-based data and a keen interest in trialing these models by every healthcare specialty.

This review aims to provide a comprehensive evaluation of publications related to AI applications in healthcare in 2024 and a comparative analysis of the trends for the past few years. We continue to use maturity-based assessments of the publications to sift through high volumes of research, and use a deep learning model, which performs with a high degree of accuracy [4]. Once again, we provide a manual analysis of data and model type to provide a detailed overview of the selected publications. With the growth in the use of generative AI, we continue to provide quantitative analysis and a review of publications related to the newly evolving field of generative AI in healthcare.

## METHODOLOGY

We performed a PubMed search (**Figure 1**) using the terms, “machine learning” or “artificial intelligence” and “2024”, restricted to English language and human subject research as of December 31, 2024 on January 1st 2025. This search resulted in an initial pool of 28,180 publications. Our methodology has remained consistent since 2019, which allows for comparative analysis of publications for each medical specialty, year over year [2,3]

**Figure 1:**
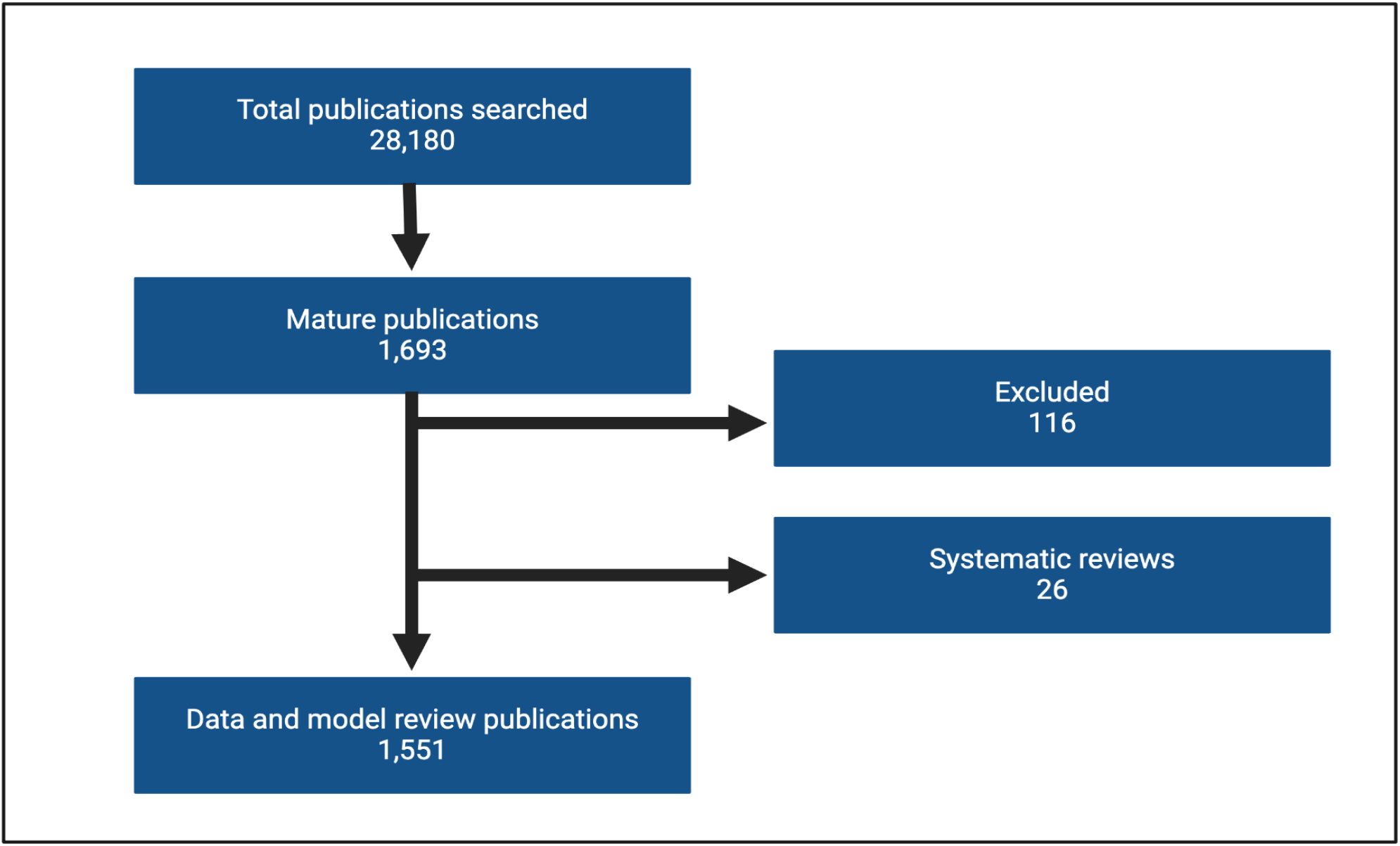
Flowchart depicting the screening and data extraction of publications

We performed a qualitative evaluation of the publications’ maturity with additional details related to the type of data used and the type of models developed across the healthcare spectrum. We used a Bidirectional Encoder Representation from Transformer (BERT)-based maturity classification model that was pre-trained and validated on manually labeled data for ‘Mature’ and ‘Not Mature’ publications [4], to assess the level of maturity of each publication. The level of maturity was determined by the ability of the publication to answer the question: “Does the output of the proposed model have a direct, actionable impact on patient care by providing information to healthcare providers or automated systems?” Systematic reviews were excluded from the count of “mature” publications as they do not independently address the above question, as the models are variable in the systematic reviews [4].

Identified publications were manually reviewed and 116 publications were excluded from the 1612 mature ones. Most of these were related to robotic surgeries or non-human studies. Further mature publications were classified based on the healthcare specialty. The General category contains many of the publications related to general AI topics that were not specialty-specific, for instance drug development-related publications. Review articles such as systematic reviews or scoping reviews were separately classified and removed prior to performing further data and model-type analysis. Education and administrative aspects of healthcare-related publications are classified as a separate specialty to provide focused reviews for those groups of publications.

Lastly, similar to prior years, we manually annotated specific details from the remaining mature publications, such as data type & model type [2]. Data type was classified manually into four categories of data: image, text, tabular, and voice. Few publications used more than one type of data, for which credit was given to each of the data categories. Model type was also manually curated from abstracts into eight different categories: Deep Learning (DL), Machine Learning (ML), AI General, Statistical, Natural Language Processing (NLP), Large Language Model (LLM), Large Vision Model (FM-LVM), and Multimodal Model (FM-MM). Those publications where the model type was not described in the title or the abstract were placed in the “AI General” class. Foundation models were separately classified as Large Language Model (LLM), Large Vision Model (FM-LVM), and Multimodal Model (FM-MM). Many of these publications were related to validation studies related to proprietary AI models. Similarly, studies using statistical analysis to analyze surveys or performance of AI models were included in the statistical class. Due to the significant increase in LLM publications in 2024, we report and provide an analysis of only mature publications using foundation models including LLMs. This is different from last year where we did a separate search for LLM in healthcare publications and provided an analysis.[2].

## RESULTS

The number of publications continues to increase as depicted by the number of articles identified by our standardized search methodology, reaching 28,180 in the year 2024. **(Figure 2A)**. Qualitatively assessed for maturity, the number of publications curated through this method also saw a significant increase from the prior year (n = 1693) [2] **(Figure 2B)**.

**Figure 2:**
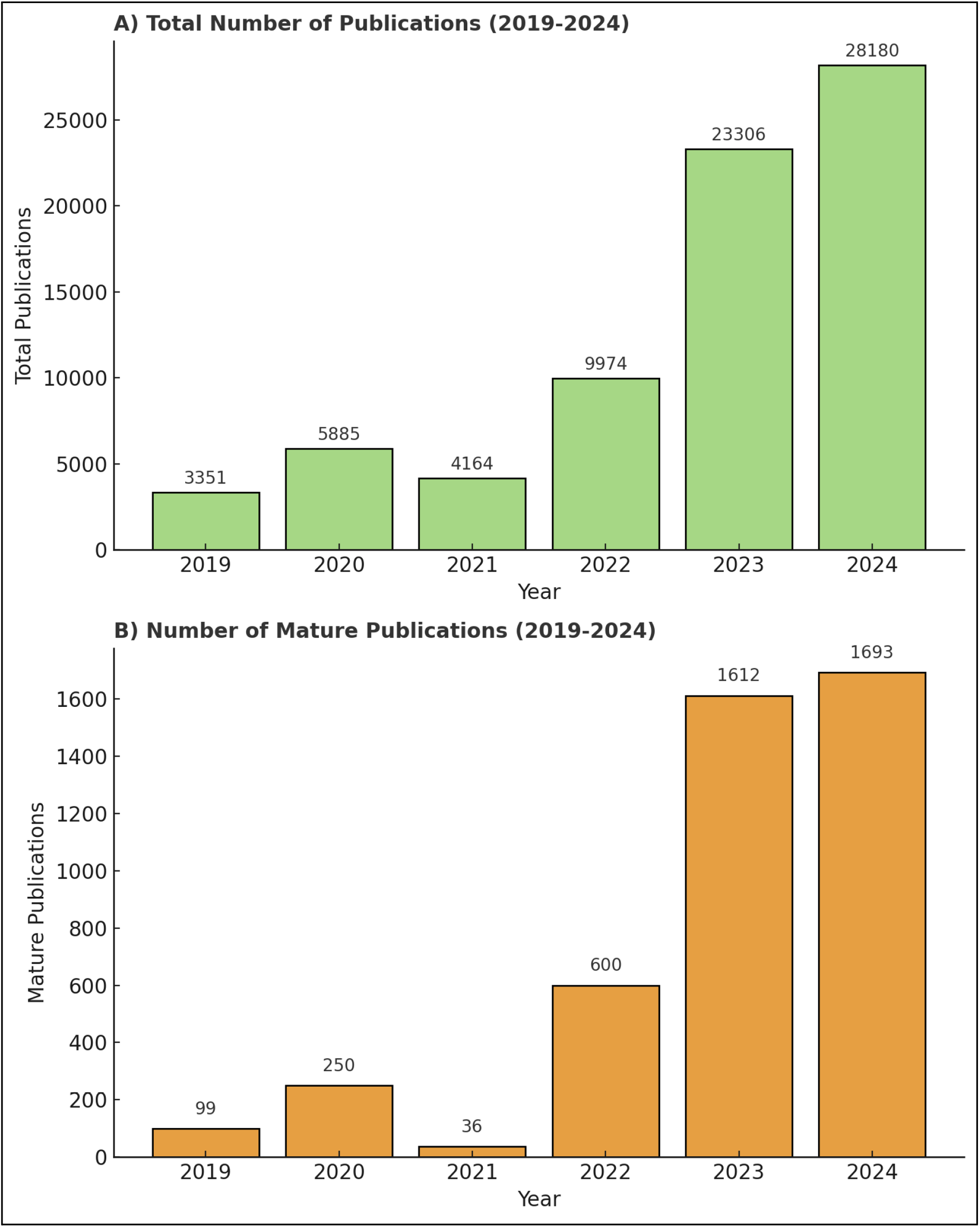
A) Bar plot showing the total number of publications from 2019-2024. B) Bar plot showing the number of mature publications from 2019-2024.

**Figure 3:**
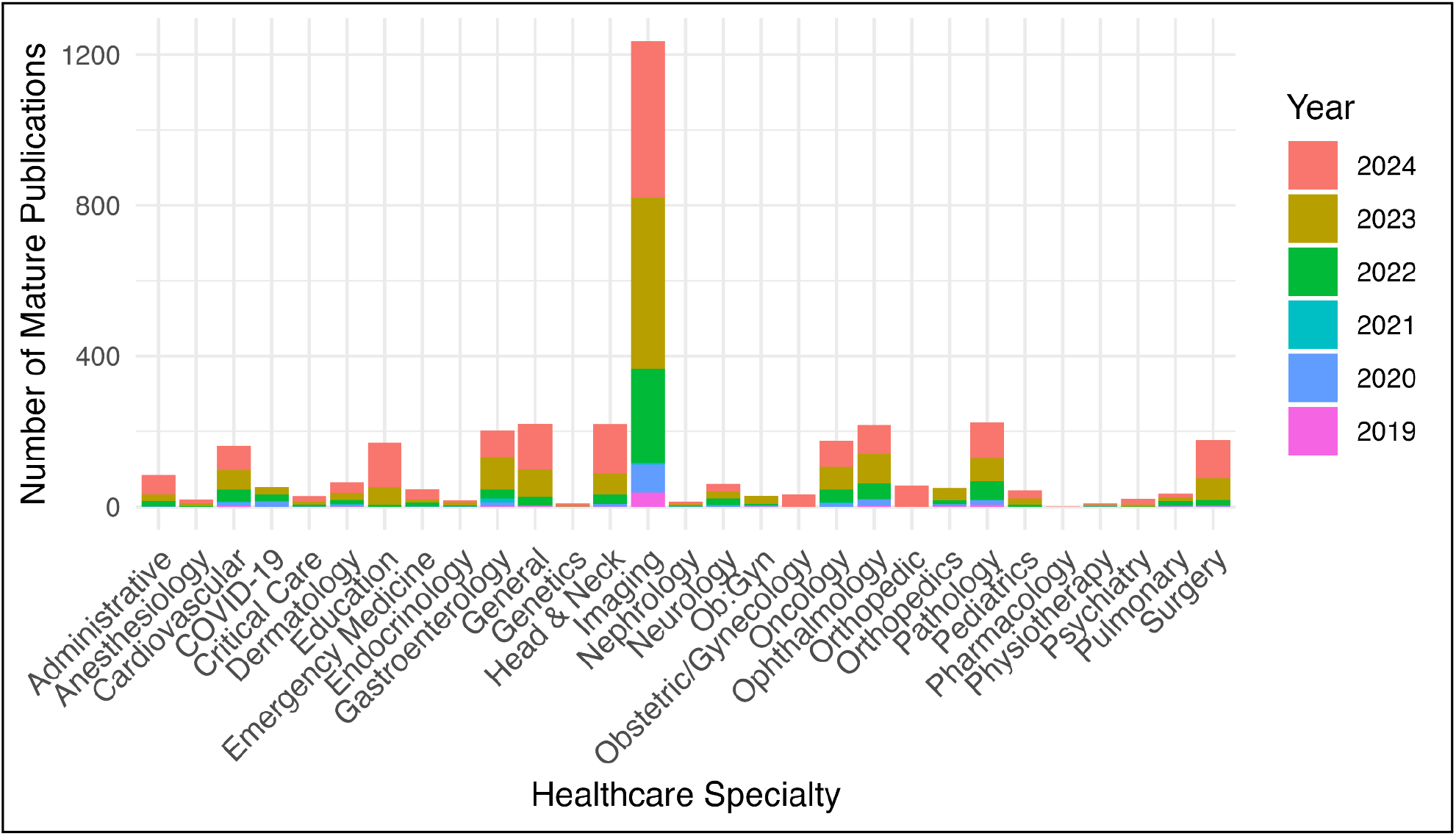
Five-year distribution in the number of mature publications in different healthcare specialties

The distribution of mature articles across specialties was not evenly distributed, led by Imaging with 407 publications, followed by Head & Neck (includes dental) with 127, and General with 122 publications in the top 3. We have observed a significant number of publications dedicated to Education that primarily use Large Language Models. **(Table 1)**.

**Table 1.** Number of mature publications by healthcare specialty for the year 2024.

| Specialty | Number of Mature Publications (2024) |
| --- | --- |
| Imaging | 407 |
| Head and Neck | 127 |
| General | 122 |
| Education | 115 |
| Surgery | 102 |
| Pathology | 95 |
| Ophthalmology | 75 |
| Gastroenterology | 70 |
| Oncology | 69 |
| Cardiology | 62 |
| Orthopedics | 54 |
| Administrative | 52 |
| Obstetric/Gynecology | 33 |
| Dermatology | 29 |
| Emergency Medicine | 28 |
| Pediatrics | 21 |
| Neurology | 20 |
| Critical Care | 15 |
| Psychiatry | 15 |
| Anesthesiology | 10 |
| Pulmonary | 9 |
| Nephrology | 7 |
| Endocrinology | 5 |
| Genetics | 4 |
| Rehabilitation Medicine | 3 |
| Pharmacology | 1 |
| COVID-19 | 1 |

Overall, Imaging continues to be the specialty with the highest number of mature articles, followed by Head & Neck, and General. Image-based specialties such as Gastroenterology, Oncology, Pathology, Ophthalmology & Orthopedics continue to dominate in the quantity of mature publications. Education, which saw no mature publications until 2021 as a separate specialty category, had substantial mature publications primarily related to the trials of LLM in these areas. In contrast, specialties such as Anesthesiology, Pulmonary, Genetics, Nephrology, and Psychiatry had fewer mature publications, with most of them using tabular or text data.

We evaluated the distribution of AI models used across all publications. LLMs have, for the first time, emerged as the most prominent with 479 publications, followed by AI General with 448 publications, DL (Deep Learning) with 372, ML (Machine Learning) with 205, NLP (Natural Language Processing) with 12, LVM (Large Vision Model) with 4, and statistical approaches with 15 publications **(Table 2)**. For the first time, we have seen the use of multimodal foundation models (FM-MM) in healthcare publications (25 publications). We also found the use of different model types in various combinations, ML and DL (7 publications), ML and NLP (2 publications), and DL and NLP (1 publication).

**Table 2:** Distribution of model types utilized in the mature articles in the year 2024 (DL = Deep Learning, ML = Machine Learning, LLM = Large Language Model, NLP = Natural Language Processing, FM-LVM = Large Vision Model, FM-MM = Multimodal model)

| Model type | Number of mature publications |
| --- | --- |
| LLM | 479 |
| AI General | 448 |
| DL | 372 |
| ML | 205 |
| FM-MM | 25 |
| Statistical | 15 |
| NLP | 12 |
| FM-LVM | 4 |

We analyzed the distribution of data types across all mature publications, of which image data remains the dominant type in 903 publications. Text data has seen a substantial increase due to the substantial increase in research using LLMs, with its use in 525 publications this year. Tabular data was utilized in 149 publications, and audio data in only 8 publications **(Figure 4)**. It is important to note that 45 publications used more than one type of data, for which credit was given to each of the data categories.

**Figure 4:**
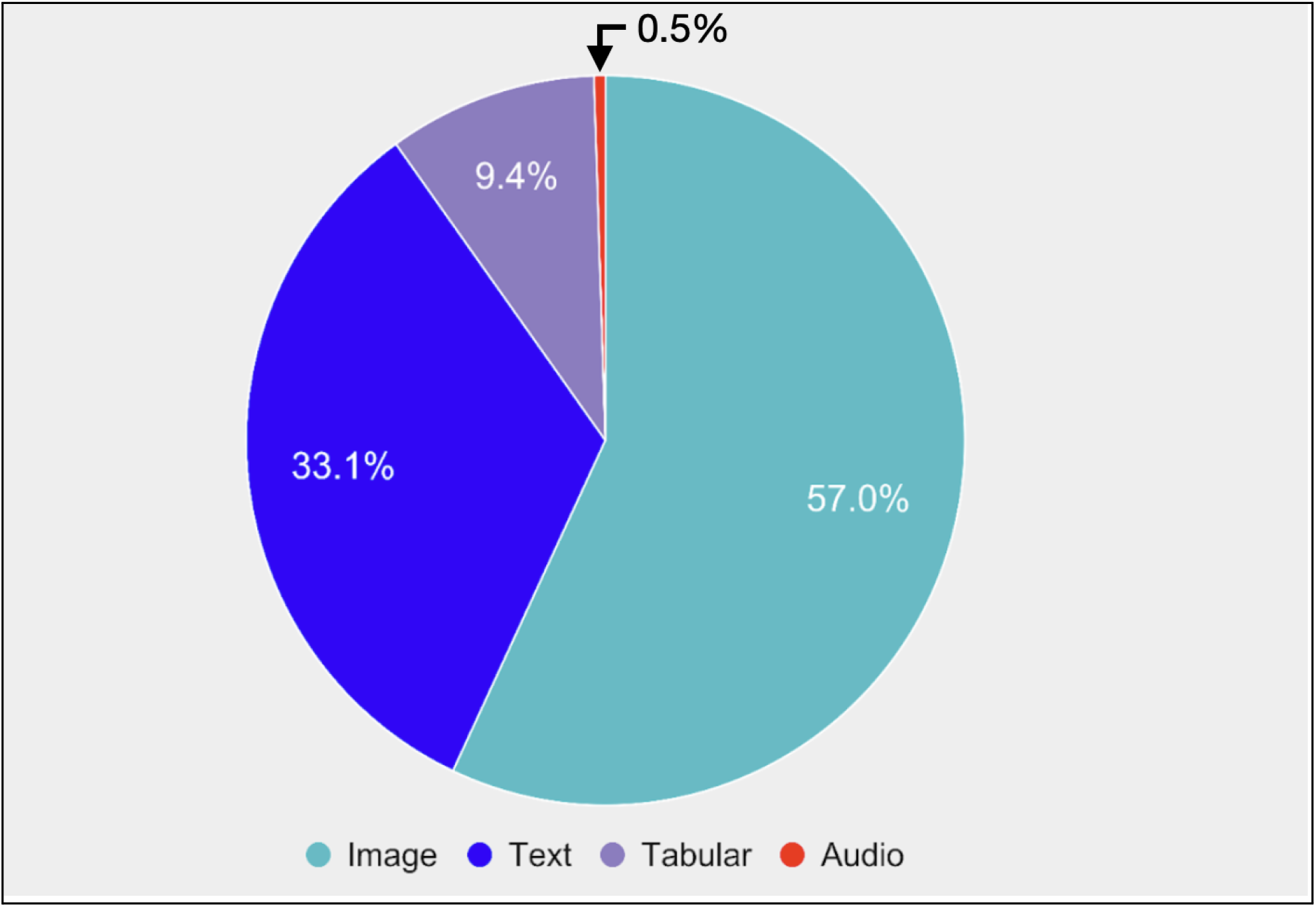
Distribution of data types utilized in the mature articles in the year 2024

Foundation model-related publications have seen a substantial increase, including those using LLMs, multimodal models, and large vision models. Education (89) and administrative (32) applications of LLM research have seen substantial research interest and publications. Surgical specialties such as Head and Neck (52), Surgery (48), and Orthopedics are also among the top healthcare specialties researching the use of LLM building on their experiences from the use of deep learning methods and early adoption of AI research. Oncology (29) and Imaging (26), which have dominated AI-based research in the past many years, are included in the top few specialties trialing foundation models, with Imaging especially experimenting with multimodal models. Specialties such as Gastroenterology (5), Pathology (10), and Cardiology (8) which have also led in AI-based research and publications for the past many years, had fewer LLM-based publications, Anesthesiology (3), Nephrology (4), and Pulmonary (1) as healthcare specialties, similar to overall AI trends have an opportunity to learn to increase the levels of research on large language models **(Figure 5)**.

**Figure 5:**
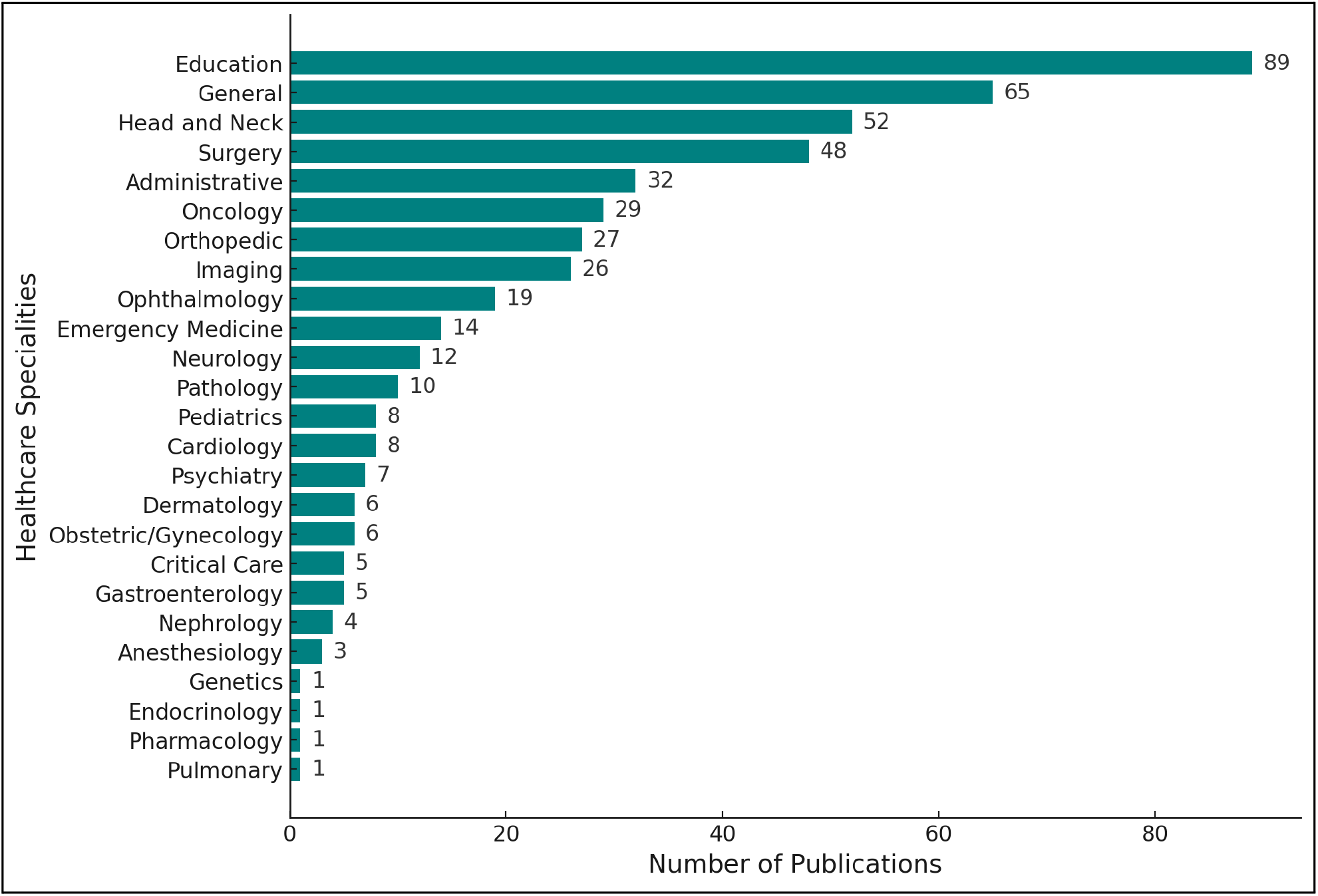
Bar plot of the number of publications based on Large Language Models across healthcare specialties

## DISCUSSION

AI in healthcare publications continues to see sustained increases in both the quantity 28,180) and quality (1793 [6.0%] mature) of publications. We observed a decrease in the number of exclusions, both due to errors related to PubMed search results and based on the number of scoping or systematic reviews. In 2024, once again, LLM-based publications continue to increase substantially across all healthcare specialties. Multimodal foundation models for the first time were experimented with, moving beyond LLMs and thereby bringing into focus many of the data types used in combination. Head & Neck (includes dental) publications experimenting with both deep learning techniques on image data and the use of LLMs saw a substantial increase relative to other publications. A substantial increase in text data for research is also due to increased focus on LLM-based research, a model type that, for the first time, had the highest number of publications. The use of proprietary models, noted as AI General, for trials is exciting to see and demonstrates the maturity of the field, moving from the development of models and internal validations to external validation trials.

Image-based AI applications in healthcare specialties such as Head & Neck, Gastroenterology, Ophthalmology, Oncology, Surgery, Head & Neck, and General categories continue to dominate in the number of mature publications (**Table 1**). These healthcare specialties, which have been leading in research and publications related to AI in healthcare, have also seen significant adoption of foundation models for experimentation. Another interesting trend is in the research related to the application of AI, especially LLMs, in areas of healthcare such as education and administrative tasks (**Figure 5**). Anesthesiology, Critical Care, Nephrology, and Rehabilitation continue to have low volumes of publications as they lack image-based data for research and have probably not developed resources for training, research and development similar to the leading healthcare subspecialties(**Table 1**). Not unexpectedly, we now observe a significant drop in publications related to COVID-19, which during the pandemic years had to be defined as a specialty of its own.

Maturity amongst Imaging as a healthcare specialty is reflected in external validations, and multicenter trials of many of the proprietary commercial AI models, moving beyond internal validations [5,6]. Many of their publications also focus on the impact of the implementation of these models both on the radiologists’ workflow and patient perceptions of their application [7,8]. Similarly, many of the publications in Gastroenterology focused on endoscopic images, are focused on trials of commercially available and device-implemented AI models undergoing multicenter trials [9–12]. Automation of Echocardiography, for ventricular function assessment, is a dominant area of study in Cardiology as a healthcare specialty [13,14]. Surgical publications, including those for Head and Neck, include a significant number of dentistry-focused publications, and have focused on the interpretation of images for decision-making [15–17]. Oncology-based publications feature the use of AI in various cancer screenings, such as for breast, cervical, and prostate cancer [18–20]. Additionally, many of the oncology publications focused on radiotherapy planning using cancer images [21]. Many other specialties, such as Ophthalmology and Neurology, are advancing from early phases of AI model development and trials to external validation studies to study real-world outcomes of their implementation [22,23]. Democratizing key healthcare clinician skill sets is an important aspect of value-based implementation of AI and is demonstrated in the use of estimation of gestational age from blind ultrasound sweeps, making care possibly available to patients with limited access to healthcare [24].

For the first time, we see LLMs and the AI General categories as the dominant categories of models in research, overtaking deep learning models. However, many of the proprietary models that have not been described in detail in the title and abstract of the publications amongst the AI General category might be utilizing deep learning models. The use of proprietary models for external validations also demonstrates and validates the maturity of AI applications being researched in healthcare, with over a thousand FDA-approved AI/ML devices [25]. There continues to be a significant increase in the utilization of text data was also observed, which may be attributed to the rapid advancement of large language models (LLMs). This advancement was evident in the distribution of model types across all papers, with LLMs being utilized in (479 [30.7 %]) of the analyzed publications. As predicted in last year’s review for the first time, we have also witnessed research related to multimodal foundation models that combine a variety of data types in healthcare [2]. We anticipate the use of these models to only grow with the use of combined data types and agent-based research in the near future.

Research related to the use of foundation models has seen exponential growth over the last two years. While LLMs have been the dominant models, large vision models and multimodal models that have been recently introduced are also being studied for application in healthcare [26–28]. These trials have a dominant focus on educational and administrative aspects of healthcare across many specialties. Publications related to the comparison between clinician experts’ performance on proficiency tests such as board examinations remain significant [29–31]. The use of LLMs for scholarly activity, framing of questions for trainee examinations, evaluation of student performance, and research applications such as the conduct of peer review has also gained significant interest [32–35]. Addressing patient queries, providing readable and understandable healthcare content for informed consent, and patient education are additional areas of interest across most of the healthcare specialties [36,37]. Unburdening the clinicians by addressing the administrative aspects of healthcare using LLMs is another area of research where studies related to the automation of discharge summaries, responses to patient messages, and generation of billing codes continue to grow [38–40]. GPT continues to be the dominant model for comparison, but many other models are also being trialed and compared for their accuracy and efficacy.

We acknowledge certain limitations of our analysis based on our search or analysis methodology. The search was limited to PubMed and with the restrictions mentioned in the methodology section. It is possible that some of the significant publications might have been missed due to our methodology. In prior years, we have manually excluded many publications from the PubMed search and classified publications into various select healthcare specialities. Due to the volume of publications (28,180), it was not possible to perform any manual evaluation of all the publications. We have tried various methods, including BERT and LLMs, for automated classification, including fine-tuning on prior data, but the results were not satisfactory. Analysis of mature publications provides a more focused review, and a consistent methodology also allows us to provide year-over-year comparative evaluations [2,3]. We have also evaluated alternate approaches to learn maturity, specifically focusing on methodologies derived from scholarly literature and journal metrics, such as no. of citations, journal impact factors, and journal H-index etc. However, it is important to note that using these metrics to evaluate maturity can be misleading when applied to the analysis of articles from the most recent year due to the unpredictable dynamic trends of these metrics [41–43]. The current BERT-based approach, despite its limitations, remains content-aware, reproducible even with changing factors such as article citation count and journal impact factor, and fine-tunable with new data.

## CONCLUSION

The maturity of the research publications related to AI in healthcare is evident from the increasing external validation trials of proprietary models across many of the leading healthcare specialties. The application of foundation models in healthcare-related publications is transforming the capabilities of AI with the increasing use of text and combined modalities of data, especially in non-clinical areas such as education and administrative aspects of healthcare. The quantity and, more importantly, the maturing quality of AI research in healthcare continue to grow across all healthcare specialties.

## DATA AVAILABILITY

Data is available upon request via email to the corresponding author or by contacting us through our website, BrainXAI Research (https://www.brainxai.com/research). Additional related publications and datasets related to AI in Healthcare can be accessed from BrainX Community website(https://www.brainxai.org).

## Notes

### Competing Interest Statement

The authors have declared no competing interest.

### Funding Statement

This study did not receive any funding

### Summary of Updates

We have just made some punctuation changes and stylistic changes in the abstract in the abstract for ease of read.

## REFERENCES

1. De Angelis L, Baglivo F, Arzilli G, Privitera GP, Ferragina P, Tozzi AE, et al. ChatGPT and the rise of large language models: the new AI-driven infodemic threat in public health. Frontiers in public health. 2023;11. doi:10.3389/fpubh.2023.1166120

2. Awasthi R, Mishra S, Grasfield R, et al. Artificial Intelligence in Healthcare: 2023 Year in Review. medRxiv. 2024:2024.2002.2028.24303482.

3. Awasthi R, Mishra S, Cywinski JB, Maheshwari K, Khanna AK, Papay FA, et al. Quantitative and Qualitative evaluation of the recent Artificial Intelligence in Healthcare publications using Deep-Learning. medRxiv. 2023. p. 2022.12.31.22284092. doi:10.1101/2022.12.31.22284092

4. Zhang J, Whebell S, Gallifant J, Budhdeo S, Mattie H, Lertvittayakumjorn P, et al. An interactive dashboard to track themes, development maturity, and global equity in clinical artificial intelligence research. The Lancet Digital health. 2022;4. doi:10.1016/S2589-7500(22)00032-2

5. van Leeuwen KG, Schalekamp S, Rutten MJCM, Huisman M, Schaefer-Prokop CM, de Rooij M, van Ginneken B, Maresch B, Geurts BHJ, van Dijke CF, Laupman-Koedam E, Hulleman EV, Verhoeff EL, Meys EMJ, Mohamed Hoesein FAA, Ter Brugge FM, van Hoorn F, van der Wel F, van den Berk IAH, Luyendijk JM, Meakin J, Habets J, Verbeke JIML, Nederend J, Meys KME, Deden LN, Langezaal LCM, Nasrollah M, Meij M, Boomsma MF, Vermeulen M, Vestering MM, Vijlbrief O, Algra P, Algra S, Bollen SM, Samson T, von Brucken Fock YHG; Project AIR Working Group. Comparison of Commercial AI Software Performance for Radiograph Lung Nodule Detection and Bone Age Prediction. Radiology. 2024 Jan;310(1):e230981. doi: 10.1148/radiol.230981. PMID: 38193833.

6. Postiglione TJ, Guillo E, Heraud A, Rossillon A, Bartoli M, Herpe G, Adam C, Fabre D, Ardon R, Azarine A, Haulon S. Multicentric clinical evaluation of a computed tomography-based fully automated deep neural network for aortic maximum diameter and volumetric measurements. J Vasc Surg. 2024 Jun;79(6):1390-1400.e8. doi: 10.1016/j.jvs.2024.01.214. Epub 2024 Feb 5. PMID: 38325564.

7. Yoon SH, Park S, Jang S, Kim J, Lee KW, Lee W, Lee S, Yun G, Lee KH. Use of artificial intelligence in triaging of chest radiographs to reduce radiologists’ workload. Eur Radiol. 2024 Feb;34(2):1094–1103. doi: 10.1007/s00330-023-10124-1. Epub 2023 Aug 24. PMID: 37615766.

8. Viberg Johansson J, Dembrower K, Strand F, Grauman Å. Women’s perceptions and attitudes towards the use of AI in mammography in Sweden: a qualitative interview study. BMJ Open. 2024 Feb 14;14(2):e084014. doi: 10.1136/bmjopen-2024-084014. PMID: 38355190; PMCID: PMC10868248.

9. Zou PY, Zhu JR, Zhao Z, Mei H, Zhao JT, Sun WJ, Wang GH, Chen DF, Fan LL, Lan CH. Development and application of an artificial intelligence-assisted endoscopy system for diagnosis of Helicobacter pylori infection: a multicenter randomized controlled study. BMC Gastroenterol. 2024 Sep 30;24(1):335. doi: 10.1186/s12876-024-03389-3. PMID: 39350033; PMCID: PMC11440712.

10. A newly developed deep learning-based system for automatic detection and classification of small bowel lesions during double-balloon enteroscopy examination. 10.1186/s12876-023-03067-w

11. Yuan XL, Liu W, Lin YX, Deng QY, Gao YP, Wan L, Zhang B, Zhang T, Zhang WH, Bi XG, Yang GD, Zhu BH, Zhang F, Qin XB, Pan F, Zeng XH, Chaudhry H, Pang MY, Yang J, Zhang JY, Hu B. Effect of an artificial intelligence-assisted system on endoscopic diagnosis of superficial oesophageal squamous cell carcinoma and precancerous lesions: a multicentre, tandem, double-blind, randomised controlled trial. Lancet Gastroenterol Hepatol. 2024 Jan;9(1):34–44. doi: 10.1016/S2468-1253(23)00276-5. Epub 2023 Nov 10. Erratum in: Lancet Gastroenterol Hepatol. 2024 Jul;9(7):e10. doi: 10.1016/S2468-1253(24)00168-7. PMID: 37952555.

12. Spada C, Piccirelli S, Hassan C, Ferrari C, Toth E, González-Suárez B, Keuchel M, McAlindon M, Finta Á, Rosztóczy A, Dray X, Salvi D, Riccioni ME, Benamouzig R, Chattree A, Humphries A, Saurin JC, Despott EJ, Murino A, Johansson GW, Giordano A, Baltes P, Sidhu R, Szalai M, Helle K, Nemeth A, Nowak T, Lin R, Costamagna G. AI-assisted capsule endoscopy reading in suspected small bowel bleeding: a multicentre prospective study. Lancet Digit Health. 2024 May;6(5):e345–e353. doi: 10.1016/S2589-7500(24)00048-7. PMID: 38670743.

13. Sveric KM, Ulbrich S, Dindane Z, Winkler A, Botan R, Mierke J, Trausch A, Heidrich F, Linke A. Improved assessment of left ventricular ejection fraction using artificial intelligence in echocardiography: A comparative analysis with cardiac magnetic resonance imaging. Int J Cardiol. 2024 Jan 1;394:131383. doi: 10.1016/j.ijcard.2023.131383. Epub 2023 Sep 26. PMID: 37757986.

14. Stowell CC, Howard JP, Ng T, Cole GD, Bhattacharyya S, Sehmi J, Alzetani M, Demetrescu CD, Hartley A, Singh A, Ghosh A, Vimalesvaran K, Mangion K, Rajani R, Rana BS, Zolgharni M, Francis DP, Shun-Shin MJ. 2-Dimensional Echocardiographic Global Longitudinal Strain With Artificial Intelligence Using Open Data From a UK-Wide Collaborative. JACC Cardiovasc Imaging. 2024 Aug;17(8):865–876. doi: 10.1016/j.jcmg.2024.04.017. Epub 2024 Jul 10. PMID: 39001730.

15. Zhao T, Wu H, Leng D, Yao E, Gu S, Yao M, Zhang Q, Wang T, Wu D, Xie L. An artificial intelligence grading system of apical periodontitis in cone-beam computed tomography data. Dentomaxillofac Radiol. 2024 Oct 1;53(7):447–458. doi: 10.1093/dmfr/twae029. PMID: 38960866.

16. Zhang JW, Fan J, Zhao FB, Ma B, Shen XQ, Geng YM. Diagnostic accuracy of artificial intelligence-assisted caries detection: a clinical evaluation. BMC Oral Health. 2024 Sep 16;24(1):1095. doi: 10.1186/s12903-024-04847-w. PMID: 39285427; PMCID: PMC11406783.

17. Yumii K, Ueda T, Kawahara D, Chikuie N, Taruya T, Hamamoto T, Takeno S. Artificial intelligence-based diagnosis of the depth of laryngopharyngeal cancer. Auris Nasus Larynx. 2024 Apr;51(2):417–424. doi: 10.1016/j.anl.2023.09.001. Epub 2023 Oct 12. PMID: 37838567.

18. Wu J, Li R, Gan J, Zheng Q, Wang G, Tao W, Yang M, Li W, Ji G, Li W. Application of artificial intelligence in lung cancer screening: A real-world study in a Chinese physical examination population. Thorac Cancer. 2024 Oct;15(28):2061–2072. doi: 10.1111/1759-7714.15428. Epub 2024 Aug 29. PMID: 39206529; PMCID: PMC11444925.

19. Winkler JK, Kommoss KS, Toberer F, Enk A, Maul LV, Navarini AA, Hudson J, Salerni G, Rosenberger A, Haenssle HA. Performance of an automated total body mapping algorithm to detect melanocytic lesions of clinical relevance. Eur J Cancer. 2024 May;202:114026. doi: 10.1016/j.ejca.2024.114026. Epub 2024 Mar 19. PMID: 38547776.

20. Warner-Smith M, Ren K, Mistry C, Walton R, Roder D, Bhola N, McGill S, O’Brien TA. Protocol for evaluating the fitness for purpose of an artificial intelligence product for radiology reporting in the BreastScreen New South Wales breast cancer screening programme. BMJ Open. 2024 May 28;14(5):e082350. doi: 10.1136/bmjopen-2023-082350. PMID: 38806433; PMCID: PMC11138303.

21. Tsang DS, Tsui G, Santiago AT, Keller H, Purdie T, Mcintosh C, Bauman G, La Macchia N, Parent A, Dama H, Ahmed S, Laperriere N, Millar BA, Liu V, Hodgson DC. A Prospective Study of Machine Learning-Assisted Radiation Therapy Planning for Patients Receiving 54 Gy to the Brain. Int J Radiat Oncol Biol Phys. 2024 Aug 1;119(5):1429–1436. doi: 10.1016/j.ijrobp.2024.02.022. Epub 2024 Mar 1. PMID: 38432285.

22. Wang Y, Han X, Li C, Luo L, Yin Q, Zhang J, Peng G, Shi D, He M. Impact of Gold-Standard Label Errors on Evaluating Performance of Deep Learning Models in Diabetic Retinopathy Screening: Nationwide Real-World Validation Study. J Med Internet Res. 2024 Aug 14;26:e52506. doi: 10.2196/52506. PMID: 39141915; PMCID: PMC11358665.

23. Shah S, Gonzalez Gutierrez E, Hopp JL, Wheless J, Gil-Nagel A, Krauss GL, Crone NE. Prospective multicenter study of continuous tonic-clonic seizure monitoring on Apple Watch in epilepsy monitoring units and ambulatory environments. Epilepsy Behav. 2024 Sep;158:109908. doi: 10.1016/j.yebeh.2024.109908. Epub 2024 Jul 3. PMID: 38964183.

24. Stringer JSA, Pokaprakarn T, Prieto JC, Vwalika B, Chari SV, Sindano N, Freeman BL, Sikapande B, Davis NM, Sebastião YV, Mandona NM, Stringer EM, Benabdelkader C, Mungole M, Kapilya FM, Almnini N, Diaz AN, Fecteau BA, Kosorok MR, Cole SR, Kasaro MP. Diagnostic Accuracy of an Integrated AI Tool to Estimate Gestational Age From Blind Ultrasound Sweeps. JAMA. 2024 Aug 27;332(8):649–657. doi: 10.1001/jama.2024.10770. PMID: 39088200; PMCID: PMC11350478.

25. U.S. Food and Drug Administration (FDA). Artificial intelligence and machine learning (AI/ML)-enabled medical devices. https://www.fda.gov/medical-devices/software-medical-device-samd/artificial-intelligence-and-machine-learning-aiml-enabled-medical-devices. Accessed: February 23 2024.

26. Temsah MH, Alhuzaimi AN, Almansour M, Aljamaan F, Alhasan K, Batarfi MA, Altamimi I, Alharbi A, Alsuhaibani AA, Alwakeel L, Alzahrani AA, Alsulaim KB, Jamal A, Khayat A, Alghamdi MH, Halwani R, Khan MK, Al-Eyadhy A, Nazer R. Art or Artifact: Evaluating the Accuracy, Appeal, and Educational Value of AI-Generated Imagery in DALL·E 3 for Illustrating Congenital Heart Diseases. J Med Syst. 2024 May 23;48(1):54. doi: 10.1007/s10916-024-02072-0. PMID: 38780839.

27. Roos J, Martin R, Kaczmarczyk R. Evaluating Bard Gemini Pro and GPT-4 Vision Against Student Performance in Medical Visual Question Answering: Comparative Case Study. JMIR Form Res. 2024 Dec 17;8:e57592. doi: 10.2196/57592. Erratum in: JMIR Form Res. 2025 Feb 11;9:e71664. doi: 10.2196/71664. PMID: 39714199; PMCID: PMC11683658.

28. Zhang, K., Zhou, R., Adhikarla, E. et al. A generalist vision–language foundation model for diverse biomedical tasks. Nat Med 30, 3129–3141 (2024). 10.1038/s41591-024-03185-2

29. Tran CG, Chang J, Sherman SK, De Andrade JP. Performance of ChatGPT on American Board of Surgery In-Training Examination Preparation Questions. J Surg Res. 2024 Jul;299:329–335. doi: 10.1016/j.jss.2024.04.060. Epub 2024 May 23. PMID: 38788470.

30. Tsai CY, Hsieh SJ, Huang HH, Deng JH, Huang YY, Cheng PY. Performance of ChatGPT on the Taiwan urology board examination: insights into current strengths and shortcomings. World J Urol. 2024 Apr 23;42(1):250. doi: 10.1007/s00345-024-04957-8. PMID: 38652322.

31. Tsoutsanis P, Tsoutsanis A. Evaluation of Large language model performance on the Multi-Specialty Recruitment Assessment (MSRA) exam. Comput Biol Med. 2024 Jan;168:107794. doi: 10.1016/j.compbiomed.2023.107794. Epub 2023 Nov 30. PMID: 38043471.

32. Suleiman A, von Wedel D, Munoz-Acuna R, Redaelli S, Santarisi A, Seibold EL, Ratajczak N, Kato S, Said N, Sundar E, Goodspeed V, Schaefer MS. Assessing ChatGPT’s ability to emulate human reviewers in scientific research: A descriptive and qualitative approach. Comput Methods Programs Biomed. 2024 Sep;254:108313. doi: 10.1016/j.cmpb.2024.108313. Epub 2024 Jun 28. PMID: 38954915.

33. Tarakji, Z., Kanaan, A., Saadi, S. et al. Concordance between humans and GPT-4 in appraising the methodological quality of case reports and case series using the Murad tool. BMC Med Res Methodol 24, 266 (2024). 10.1186/s12874-024-02372-6

34. Quah B, Zheng L, Sng TJH, Yong CW, Islam I. Reliability of ChatGPT in automated essay scoring for dental undergraduate examinations. BMC Med Educ. 2024 Sep 3;24(1):962. doi: 10.1186/s12909-024-05881-6. PMID: 39227811; PMCID: PMC11373238.

35. Mistry NP, Saeed H, Rafique S, Le T, Obaid H, Adams SJ. Large Language Models as Tools to Generate Radiology Board-Style Multiple-Choice Questions. Acad Radiol. 2024 Sep;31(9):3872–3878. doi: 10.1016/j.acra.2024.06.046. Epub 2024 Jul 15. PMID: 39013736.

36. Shiraishi M, Tomioka Y, Miyakuni A, Moriwaki Y, Yang R, Oba J, Okazaki M. Generating Informed Consent Documents Related to Blepharoplasty Using ChatGPT. Ophthalmic Plast Reconstr Surg. 2024 May-Jun 01;40(3):316–320. doi: 10.1097/IOP.0000000000002574. Epub 2023 Dec 19. PMID: 38133626.

37. Wu Y, Zhang Z, Dong X, Hong S, Hu Y, Liang P, Li L, Zou B, Wu X, Wang D, Chen H, Qiu H, Tang H, Kang K, Li Q, Zhai X. Evaluating the performance of the language model ChatGPT in responding to common questions of people with epilepsy. Epilepsy Behav. 2024 Feb;151:109645. doi: 10.1016/j.yebeh.2024.109645. Epub 2024 Jan 19. PMID: 38244419.

38. Schwieger A, Angst K, de Bardeci M, Burrer A, Cathomas F, Ferrea S, Grätz F, Knorr M, Kronenberg G, Spiller T, Troi D, Seifritz E, Weber S, Olbrich S. Large language models can support generation of standardized discharge summaries - A retrospective study utilizing ChatGPT-4 and electronic health records. Int J Med Inform. 2024 Dec;192:105654. doi: 10.1016/j.ijmedinf.2024.105654. Epub 2024 Oct 14. PMID: 39437512.

39. Small WR, Wiesenfeld B, Brandfield-Harvey B, Jonassen Z, Mandal S, Stevens ER, Major VJ, Lostraglio E, Szerencsy A, Jones S, Aphinyanaphongs Y, Johnson SB, Nov O, Mann D. Large Language Model-Based Responses to Patients’ In-Basket Messages. JAMA Netw Open. 2024 Jul 1;7(7):e2422399. doi: 10.1001/jamanetworkopen.2024.22399. PMID: 39012633; PMCID: PMC11252893.

40. Wang Y, Huang Y, Nimma IR, Pang S, Pang M, Cui T, Kumbhari V. Validation of GPT-4 for clinical event classification: A comparative analysis with ICD codes and human reviewers. J Gastroenterol Hepatol. 2024 Aug;39(8):1535–1543. doi: 10.1111/jgh.16561. Epub 2024 Apr 16. PMID: 38627920.

41. Waltman L, Traag VA. Use of the journal impact factor for assessing individual articles: Statistically flawed or not? F1000Res. 2020;9. doi:10.12688/f1000research.23418.2

42. Agarwal A, Durairajanayagam D, Tatagari S, Esteves SC, Harlev A, Henkel R, et al. Bibliometrics: tracking research impact by selecting the appropriate metrics. Asian J Androl. 2016;18:p 296.

43. Mustafa G, Rauf A, Ahmed B, Afzal MT, Akhunzada A, Alharthi SZ. Comprehensive Evaluation of Publication and Citation Metrics for Quantifying Scholarly Influence. [cited 27 Feb 2024]. Available: https://ieeexplore.ieee.org/abstract/document/10168127

